# Long term consequences of Total Ankle Replacement versus Ankle Fusion; a 25 year national population study of 41,000 patients

**DOI:** 10.1101/2025.02.17.25322408

**Authors:** Conor Hennessy, Simon Abram, Rick Brown, Constantinos Loizou, Robert Sharp, Adrian Kendal

**Affiliations:** Foot and Ankle Research Group Oxford, Nuffield Orthopaedic Centre, Oxford University Hospitals; Nuffield Department of Orthopaedics, Rheumatology and Musculoskeletal Sciences, University of Oxford, Oxford

## Abstract

**Aims:** Definitive and successful treatment of end stage ankle arthritis is either Total Ankle Replacement (TAR) or Ankle Fusion (AF). Both options place patients on an irreversible pathway that risks harm from further surgery. AF may predispose patients to subsequent hindfoot joint fusion and TAR is associated with high rates of complex revision surgery. The aim is to improve decision making by investigating the risks of further surgery, adjacent joint surgery and rare but serious complications of AF versus TAR.

**Methods:** An England population cohort study was performed using the Hospital Episode Statistics database, linked to ONS mortality data (19982023). The primary outcome was Kaplan Meier curve analysis of revision surgery free survival of TAR versus AF. Secondary outcome measures were the rates of adjacent joint/hindfoot fusion, any further reintervention to the ankle, perioperative mortality, 90 day complications, and serious adverse events.

**Results:** 10,335 TAR and 30,704 AF were analysed. The AF revision rate was significantly lower than TAR at all time points including; 5 years (2% vs 6.1%, RR 0.12; 95% CI 0.10 to 0.16), 10 years (2.5% vs 10.2%, RR 0.12; 95% CI 0.08 to 0.18) and 20 years (3.1% vs 13.55%, RR 0.12; 95% CI 0.01 to 0.23).

There was no significant difference in 25 year risk of adjacent joint fusion following AF (8.64%, 95% CI 7.79% to 9.58%) versus TAR (6.82%; 95% CI 5.36% to 8.66%). TAR was associated with higher risks of intra operative fracture (0.42% vs 0.10%, RR = 4.35) and reintervention for wound infection (0.26% vs 0.15%, RR 1.74) but fewer pulmonary emboli (0.23% vs 0.58%, RR = 0.40).

**Conclusion:** Both TAR and AF are safe definitive treatments of ankle arthritis with low perioperative risk. TAR is associated with a significantly higher rate of further revision surgery than AF. AF does not predispose patients to hindfoot fusion surgery.

## Introduction

Ankle arthritis is as debilitating as hip arthritis and affects approximately 1% of the global population^1,2^. In the United Kingdom, an estimated 29,000 patients present to specialist care each year with symptomatic ankle arthritis, most commonly due to post traumatic arthritis, osteoarthritis, and/or inflammatory arthropathy^3^. The two surgical options for medically intractable end stage arthritis are Total Ankle Replacement (TAR) and Ankle Fusion (AF)^4^. Both are equally successful in improving pain, function, and quality of life in the short term^5^. The choice of either TAR or AF for individual patients is based on multiple factors including the availability of outcome data. Ultimately, both of these definitive treatments irreversibly place patients on a particular path that risks the harm of further operations. We are interested in understanding the long-term risks and sequelae of these two treatments at a population level. This will better inform future decision making for patients and healthcare professionals.

Ankle fusion is a well-established treatment for advanced arthritis with reliable results and high levels of patient satisfaction^6–8^. Moreover, the development of arthroscopic ankle fusion has been shown to reduce non-union rates, length of hospital stay, intra-operative blood loss, and tourniquet time compared to open ankle fusion ^8,9^. Although superior to open AF, arthroscopic AF still has a reported non-union rate of 8-13% ^6,10–12^. Similar non-union rates were again observed in a recent single centre study of over 250 patients undergoing arthroscopic AF^13^. Importantly this longer term study used Kaplan-Meier analysis to estimate that 26% of patients require a subsequent subtalar joint (hindfoot) fusion within 9 years of AF^13^. This remains a major concern of ankle fusion surgery; that it predisposes patients to developing adjacent joint degeneration, requiring a hindfoot fusion^14^. This can shift a patient from a high functioning individual to someone with a rigid ankle-hindfoot complex ^9,15,16^.

In contrast, TAR is thought to protect against the development of adjacent joint disease by preserving a range of motion at the ankle. The major risk with TAR is the high revision rate which varies across different National Joint Registries. A recent systematic review of data from three national Registries found a revision rate of up to 20% at 5 years and a range from 32% at 10 years in Norway compared to 26% at 10 years in Sweden ^17^. There is additional variation in revision rates when comparing different TAR implants^18,19^. Failing TARs are also thought to be vulnerable to multiple interim procedures before the ultimate revision operation. These minor procedures include open/arthroscopic debridement, surgical grafting of cystic bone defects, and bearing exchange; potentially posing further harm to patients ^4,20^.

Since 2010 there has been a 77% mean increase in the number of TARs performed in the UK^21^. This equates to 974 recorded in 2019 compared to 402 in 2010. Based on these collective observations we may be facing a wave of complex, time consuming TAR revision operations which require high levels of experience, have poor outcomes and are preceded by a larger number of smaller procedures and their associated risks ^22–24^.

The randomised control trial of Total Ankle Replacement Versus Arthrodesis (TARVA) has demonstrated that TAR is as effective at improving pain and patient reported outcome measures as ankle fusion at 1 year^5^. However, such studies may not be sufficiently powered to detect rare, but serious, adverse events and they cannot readily describe the 20+ year impact of these two different surgical options. Patients with end stage ankle arthritis who are considering either of these irreversible treatments deserve to understand the long-term consequences of each choice. This study aims to interrogate national population data over a 25 year period to understand better the life time risk of further surgery and the rate of rare but serious peri-operative risks following TAR versus AF.

## Methods

We conducted an analysis of prospectively collected nationwide data from the England Hospital Episodes Statistics (HES) database, combined with Office of National Statistics (ONS) mortality data^25^. The HES database holds information on all patients admitted to NHS hospitals in England. Each record in the database relates to one finished Consultant episode, describing the time an individual spends under the care of one NHS Consultant. Procedures performed in private hospitals are excluded. Submission of records to HES is mandated for accurate remuneration of NHS hospitals, and as such HES provides universal coverage of day case and inpatient surgical care. Any procedure performed is recorded using the Office of Population Censuses and Surveys Classification of Interventions and Procedures, version 4 (OPCS-4) codes. The World Health Organization International Classification of Diseases, 10th revision (ICD-10) codes are used to record diagnoses. The database includes detailed demographic data, co-morbidities, peri-operative complications and any further operative intervention. Hospital episode statistics data were then linked to mortality records from the Office for National Statistics, which provided information about the date and cause of death.

The elective Hospital Episode Statistics database (HES) for England was interrogated to identify all patients undergoing either a total ankle replacement (TAR) or ankle fusion (AF), between 1998 and 2023. The list of OPCS-4 codes used to define procedures and re-interventions is detailed in Supplementary File 1. The AF group did not include patients who underwent tibio-talar-calcaneal (TTC) fusion or an urgent ankle fusion as part of fracture fixation. The primary outcome was the rate of revision post TAR versus AF, analysed by Kaplan-Meier revision free survival. In the case of TAR, revision surgery was defined as, and included all OPSC-4 codes pertaining to, revision arthroplasty or revision to fusion. Revision surgery post AF was defined as, and included all OPSC-4 codes pertaining to, revision of the ankle fusion Supplementary File 1. Patients were censored if they died during the study period prior to the primary outcome. Secondary outcome measures included the rates of postoperative complications at ninety days following TAR versus AF. The progression of adjacent joint disease requiring fusion was determined by the rate of hindfoot fusion and reported as hindfoot fusion free survival determined by Kaplan-Meier analysis. Hindfoot fusion included, and was defined by OPSC-4 codes for, any operative event recorded as a subtalar fusion (including talo-calcaneal fusion), talo-navicular fusion, hindfoot fusion, and/or a triple fusion. The rate, number, and type of any subsequent re-interventions following TAR or AF were compared and reported as ‘re-intervention’ free survival. This analysis included any re-intervention to the ipsilateral ankle/hindfoot post AF or TAR and was not censored on an individual basis; i.e. every re-intervention would be recorded as an event, regardless of whether it was on the same individual. Data was presented as rates of the outcome of interest with confidence intervals, and survival analysis was performed using R and StataSE.

Pearsons Chi squared tests were used to determine if the assigned intervention was associated with differences in the demographic distributions (for sex, age, IMD quintile, race, Charlson score). In order to account for the large numbers of patients, an effect estimate was used based on the Cramér’s V correction to determine if any of the statistically significant differences seen were practically significant. This method adjusts for very large population size when applying Chi squared calculations. The estimated effects following adjustment are classed as very small (Cramér’s V = 0.01-0.09), small (Cramér’s V = 0.10-0.29), medium (Cramér’s V = 0.30-0.49), large (Cramér’s V = 0.50-.69) and very large (0.70 and above).

A Cox proportional hazards model was used to calculate the adjusted Hazard Ratios of revision over time by index surgery, age group, sex, index of multiple deprivation (quintile derived from regional factors in England including average income, employment, education, housing andc rime; 1=least deprived area, 5=most deprived), race, and modified Charlson co-morbidity index score.

## Results

Following data cleaning, there were 10,335 patients that received a TAR and 30,704 patients that received an ankle fusion eligible for analysis (see Supplementary Figures 1 and 2). The mean age for TAR was 63.8 years (median = 65 years) and 55.2 years (median = 60 years) for AF. Both TAR and AF were most commonly performed in male patients (60.1% and 63.4% respectively), and in the 60-79 age group (68.74% and 48.19% respectively, Table 1).

**Table 1.** Demographic data of patients undergoing Total Ankle Replacement and Ankle fusion. Demographic data was tested to determine if differences existed in distributions based on the index intervention. Chi squared tests were used to determine whether any significant differences were present (P<0.05), and significant differences were adjusted to identify if any practical differences existed based on treatment group. * = very small (<u>Cramér’s V</u> = 0.01-0.09) #=small (<u>Cramér’s V=</u> 0.10-0.29).

|  | **Total Ankle Replacement** |  | **Ankle Fusion** |  |  |
| --- | --- | --- | --- | --- | --- |
|  | n | % | n | % | **Cramér's V** |
| *Overall* | 10,335 | 100 | 30,704 | 100 |  |
| **Sex** |  |  |  |  | **\*0.02** |
| *Male* | 6,207 | 60.06 | 19,453 | 63.36 |  |
| *Female* | 4,128 | 39.94 | 11,251 | 36.64 |  |
| **Age group** |  |  |  |  | **#0.11** |
| *0 - 19* | 20 | 0.19 | 352 | 1.15 |  |
| *20 - 39* | 148 | 1.43 | 2,859 | 9.31 |  |
| *40 - 59* | 2,112 | 20.44 | 10,916 | 35.55 |  |
| *60 - 79* | 7,104 | 68.74 | 14,796 | 48.19 |  |
| *80+* | 951 | 9.2 | 1,781 | 5.8 |  |
| **Race** |  |  |  |  | **\*0.026** |
| *White* | 10,225 | 98.94 | 29,800 | 97.06 |  |
| *Black* | 21 | 0.2 | 285 | 0.93 |  |
| *Asian* | 65 | 0.63 | 409 | 1.33 |  |
| *Mixed* | 7 | 0.07 | 76 | 0.25 |  |
| *Other* | 17 | 0.16 | 134 | 0.44 |  |
| **IMD** |  |  |  |  | **\*0.039** |
| *Most deprived 0-20%* | 1,571 | 15.2 | 6,469 | 21.07 |  |
| *More deprived 20-40%* | 1,925 | 18.63 | 6,351 | 20.68 |  |
| *Middle 40-60%* | 2,354 | 22.78 | 6,753 | 21.99 |  |
| *less deprived 60-80%* | 2,489 | 24.08 | 6,194 | 20.17 |  |
| *least deprived 80-100%* | 1,996 | 19.31 | 4,937 | 16.08 |  |
| **Charlson score** |  |  |  |  | **\*0.038** |
| *None (0)* | 5,924 | 57.49 | 19,629 | 63.93 |  |
| *Mild (1-15)* | 4,111 | 39.78 | 10,054 | 32.74 |  |
| *Moderate (16-30)* | 264 | 2.55 | 902 | 2.94 |  |
| *Severe (31-50)* | 18 | 0.17 | 119 | 0.39 |  |

Both operations were associated with low 90-day mortality rates; 0.23% for TAR and 0.41% for AF, and significantly lower 90-day mortality was observed post TAR (RR 0.57, 95% CI 0.37 to 0.89, Figure 1, and Supplementary Figure 3). Beyond seven years, the mortality was significantly greater post TAR than post AF (Supplementary Figure 3). Significantly fewer pulmonary emboli were observed within 90 days following TAR compared to AF (0.23% vs 0.58%, RR = 0.40). TAR was associated with higher risks of intra-operative fracture (0.43% vs 0.10%, RR = 4.35) and re-intervention for wound infection (0.26% vs 0.15%, RR 1.74). There was a significantly greater risk of re-intervention within 90 days for metalware complications following AF compared to TAR (2.99% AF versus 1.70% TAR, RR 0.40).

**Figure 1.**
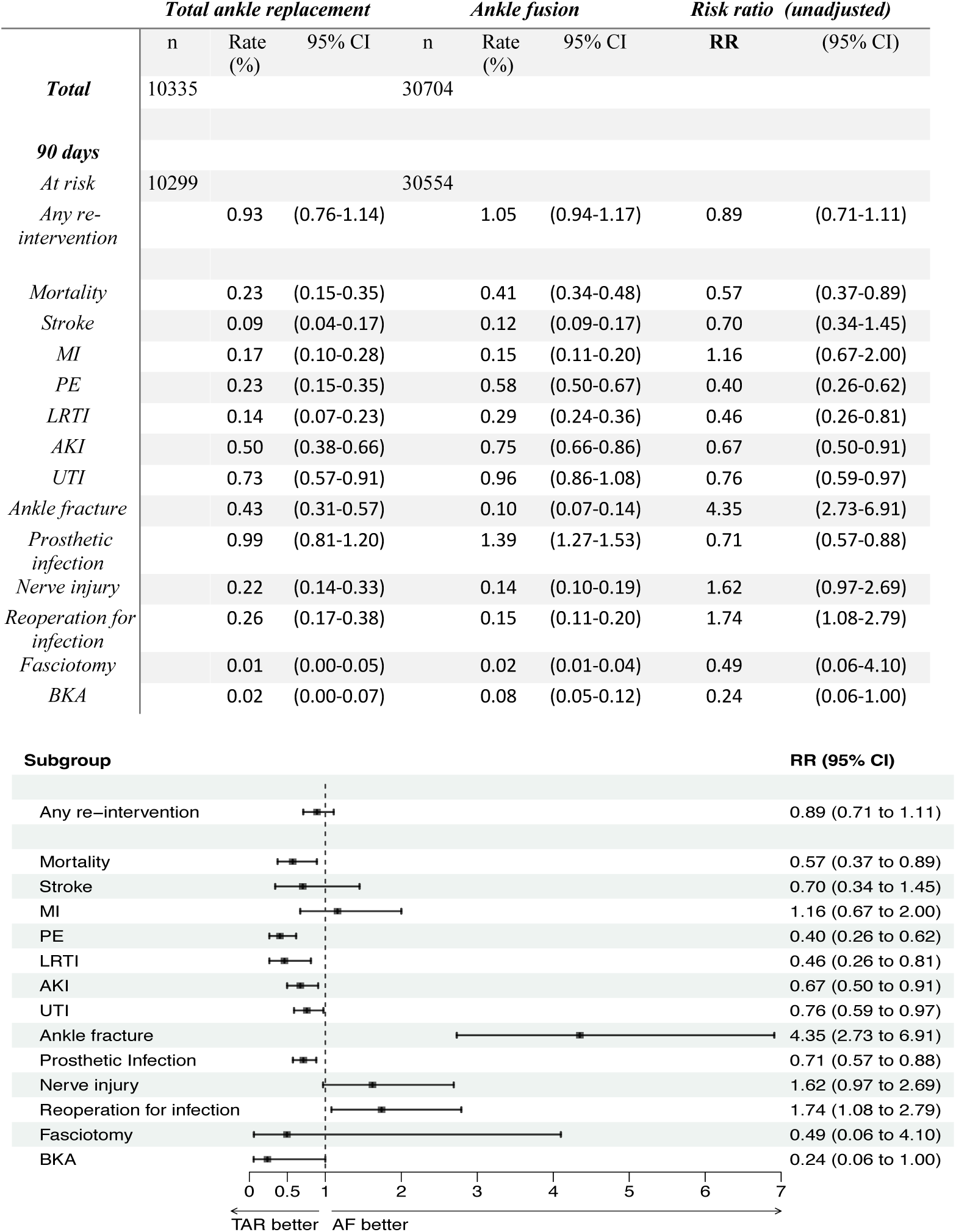
Relative Risk (RR) of 90-day complications in the Total Ankle Replacement group relative to the Ankle Fusion group. **A)** Rates of complications at 90 days are shown as absolute rate in each treatment group, with 95% CIs. Relative risk (RR) is shown in the right most column with 95% CIs. **B)** Forest plots of RR for each 90d complication outlined in the table in A.

There was a significant increase in the revision rates of TAR compared to AF at all time points (Figure 2). Revision of TAR included any, and all, OPCS-4 codes pertaining to ankle revision arthroplasty or revision to fusion. Revision of AF was defined as revision arthrodesis/fusion of ankle. The observed revision rates of TAR were 10.9% at 10 years in 4,447 patients (95% CI 9.5% to 10.9%), and 13.5% at 20 years in 661 patients (95% CI 12.6% to 14.6%). In comparison, the revision rate of post AF was 2.5% at 10 years in 13,306 patients (95% CI 2.3% to 2.7%) and 3.1% at 20 years in 1,705 patients (95% CI 2.8% to 3.4%). Thus there was an increased risk of revision in the TAR group compared to the AF group at all time points including at 10 years (RR 0.12 (95% CI 0.08 – 0.18)) and 20 years (RR (0.12 (95% CI 0.01 – 0.23)). To assess the impact of potential confounders, the adjusted hazard ratio of revision across the study period was calculated using a cox proportional hazards model. After adjusting for age, sex, index of multiple deprivation, race and Charlson co-morbidity score, TAR had a higher risk of revision (HR 3.28, 95% CI 2.92-3.67) when compared to AF (Table 2 and Figure 2B by age).

**Figure 2.**
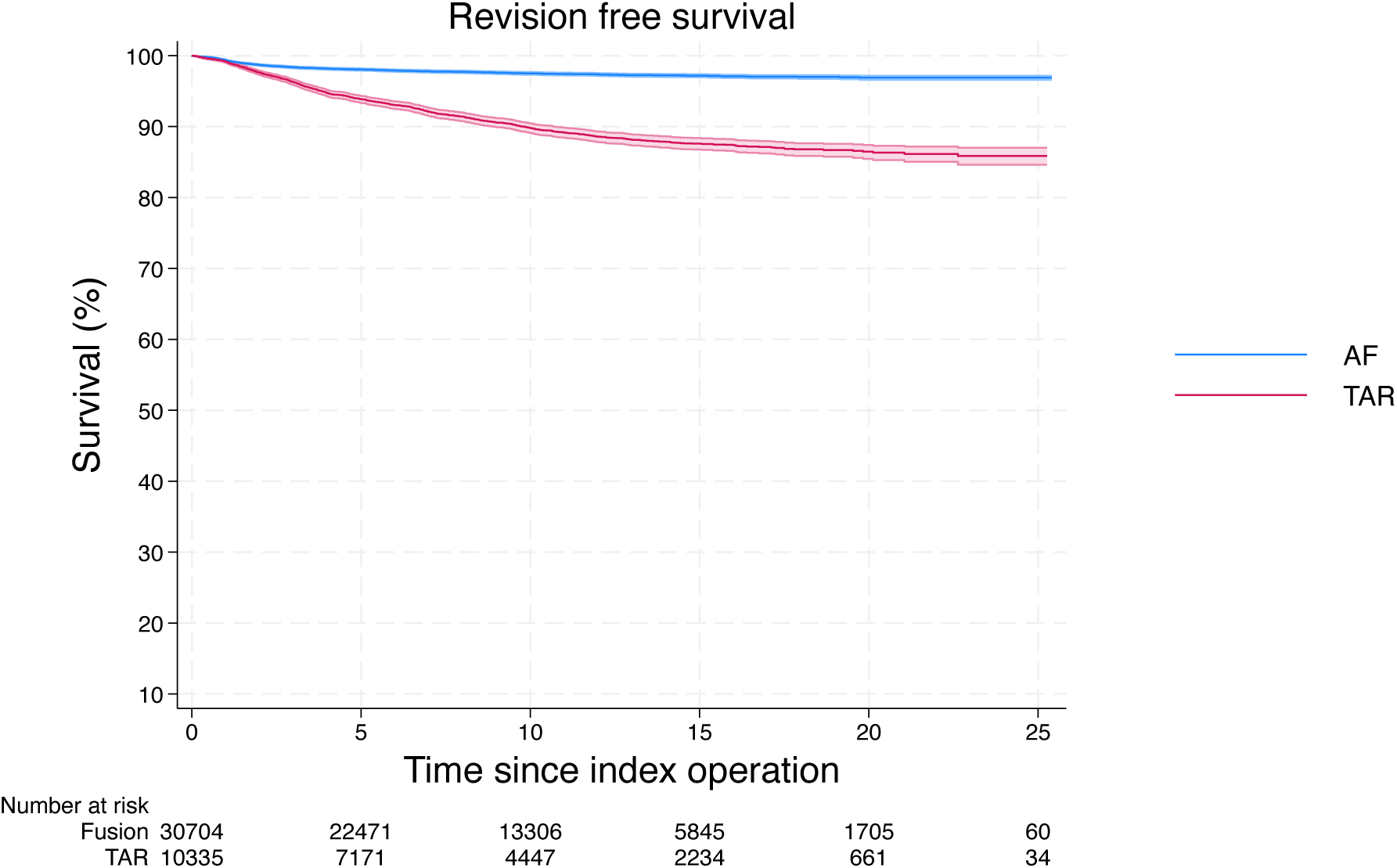
Kaplan-Meier curve of revision free survival in Total Ankle Replacement and Ankle Fusion. **A)** Death censored revision free survival over 25 years post ankle fusion (AF) and Total Ankle Replacement (TAR). Revision in TAR was classified as revision arthroplasty or revision of the TAR to a fusion. Revision in the AF group was classified as a revision of the ankle fusion.

**Table 2.** Re-interventions following Total Ankle Replacement.

| **Operation** | **Overall** |  | **0 – 90 days** |  | **90 days - 1 year** |  | **1 - 5 years** |  | **5 - 10 years** |  |
| --- | --- | --- | --- | --- | --- | --- | --- | --- | --- | --- |
|  | N | % | N | % | N | % | N | % | N | % |
| *Ankle/hindfoot Injection* | 1302 | 28.81 | 0 | 0.00 | 147 | 27.74 | 745 | 31.05 | 275 | 26.99 |
| *Explore +/- debride Ankle* | 704 | 15.50 | 21 | 16.03 | 105 | 19.81 | 411 | 17.13 | 128 | 12.56 |
| *TAR revision* | 552 | 12.18 | 13 | 9.92 | 46 | 8.68 | 267 | 11.13 | 152 | 14.92 |
| *Infection* | 426 | 9.41 | 52 | 39.69 | 53 | 10.00 | 198 | 8.25 | 82 | 8.05 |
| *Ankle aspiration* | 410 | 9.09 | 20 | 15.27 | 89 | 16.79 | 191 | 7.96 | 75 | 7.36 |
| *TAR conversion to fusion* | 306 | 6.82 | 2 | 1.53 | 22 | 4.15 | 169 | 7.04 | 90 | 8.83 |
| *Metal removal* | 241 | 5.35 | 6 | 4.58 | 29 | 5.47 | 112 | 4.67 | 68 | 6.67 |
| *Bone cyst graft* | 215 | 4.77 | 1 | 0.76 | 15 | 2.83 | 112 | 4.67 | 57 | 5.59 |
| *Hindfoot fusion* | 171 | 3.82 | 0 | 0.00 | 11 | 2.08 | 94 | 3.92 | 48 | 4.71 |
| *Ankle synovectomy* | 64 | 1.44 | 3 | 2.29 | 6 | 1.13 | 43 | 1.79 | 11 | 1.08 |
| *Amputation* | 61 | 1.34 | 2 | 1.53 | 1 | 0.19 | 28 | 1.17 | 19 | 1.86 |
| *Ankle ligament recon* | 44 | 0.97 | 7 | 5.34 | 4 | 0.75 | 17 | 0.71 | 13 | 1.28 |
| *Fracture/Fixation* | 15 | 0.32 | 3 | 2.29 | 2 | 0.38 | 6 | 0.25 | 1 | 0.10 |
| *Tendon operation* | 9 | 0.20 | 1 | 0.76 | 0 | 0.00 | 6 | 0.25 | 0 | 0.00 |
| *Total* | 4520 | 100.00 | 131 | 100.00 | 530 | 100.00 | 2399 | 100.00 | 1019 | 100.00 |

Figure 3 demonstrates the rate of subsequent hindfoot fusion following either TAR or AF. In this study, hindfoot fusion included any operative event recorded as a subtalar fusion (including talo-calcaneal fusion), talonavicular fusion, hindfoot fusion and/or a triple fusion. Over the 25-year period, 8.64% of AF patients (95% CI 7.79% to 9.58%) and 6.82% of TAR patients (95% CI 5.36% to 8.66%) required a hindfoot fusion. Thus, there was no significant difference in the risk of subsequent hindfoot fusion post TAR versus AF (RR AF vs TAR 1.26 (95% CI 0.15 – 10.28)).

**Figure 3.**
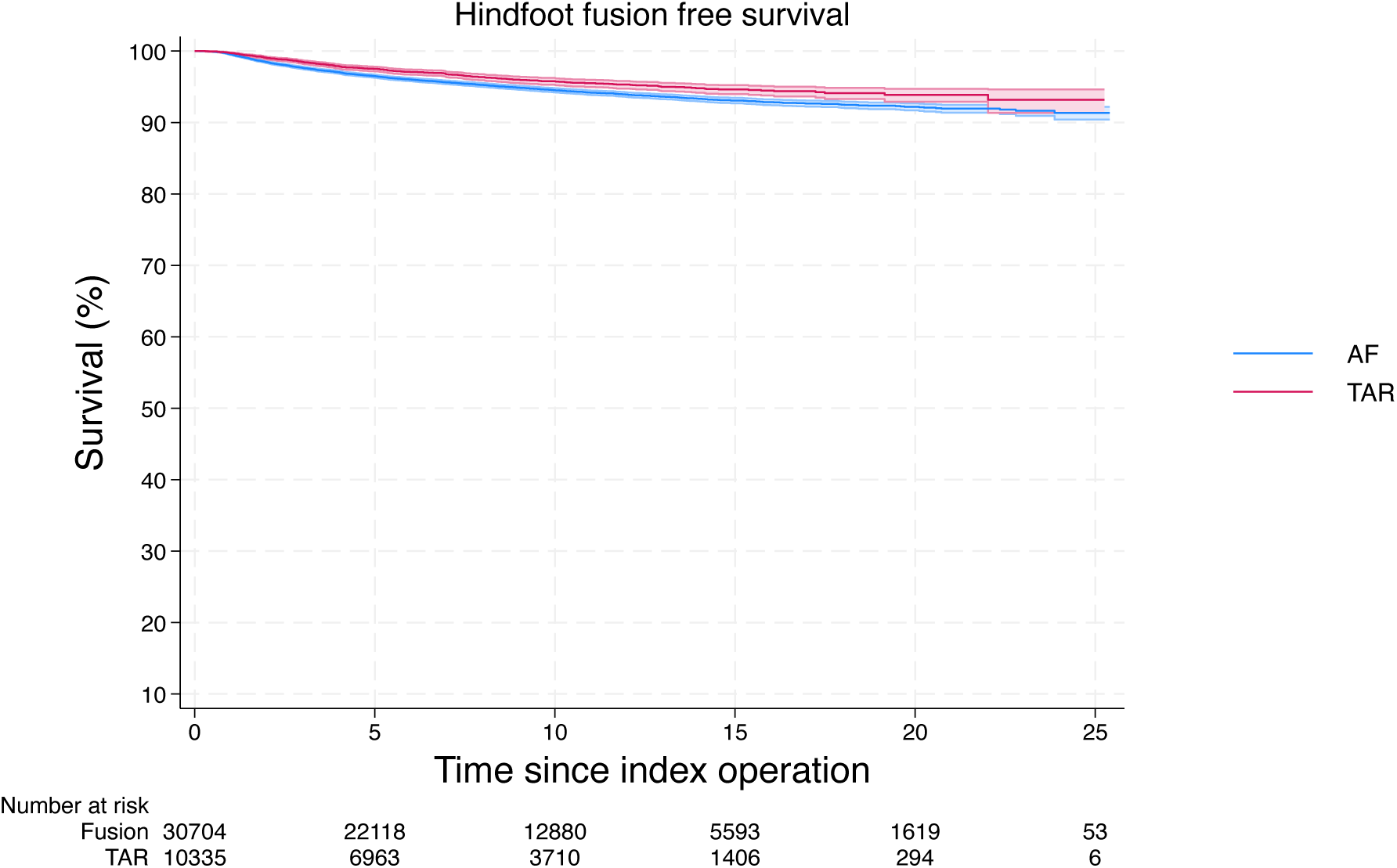
Kaplan-Meier curve of hindfoot fusion free survival in Total Ankle Replacement (TAR) and Ankle Fusion (AF)

We next explored the long-term risk of *any* further intervention to the same ankle and/or hindfoot following either TAR or AF. The overall re-intervention free survival over 25 years was 65% (95% CI 62%-68%) following TAR and 69.2% (95% CI 69%-70%) following AF (Figure 4). The commonest re-interventions after TAR were exploration/debridement of ankle joint, wound infection surgery, joint aspiration and revision TAR; together accounting for over 60% of all re-interventions at 90 days, 1 year and 5 years (Figure 5 and Table 3). There was a significant increase in the rate of re-intervention within the first year following AF (9.1%, (95% CI 8.7%-9.4%)) compared to TAR (5.0%, (95% CI 4.6% – 5.4%)). The majority of these were removal of metal, particularly in the first 90 days and 1 year following AF (Figure 6 and Table 4).

**Figure 4.**
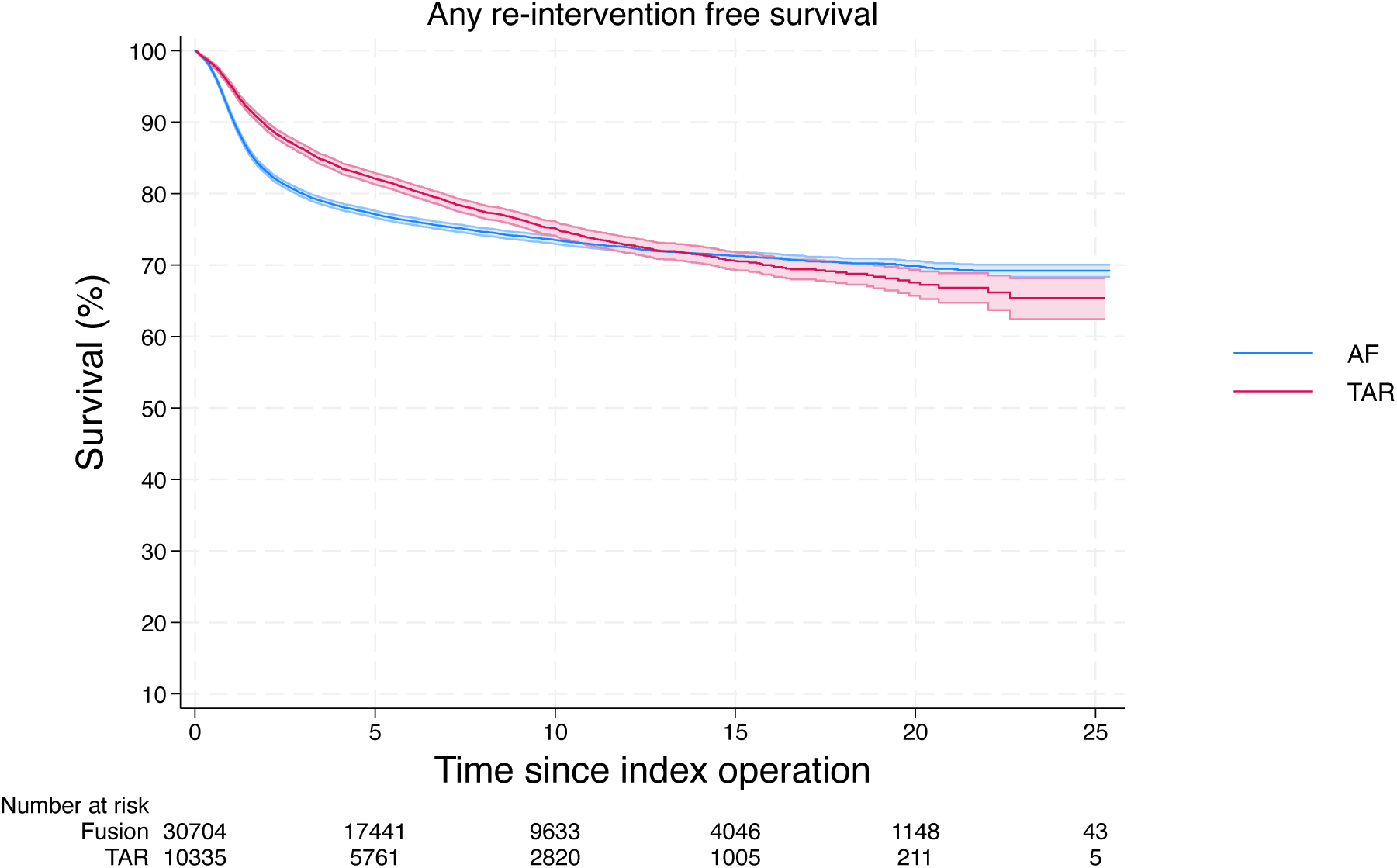
Kaplan-Meier curve of ‘any re-intervention’ free survival in TAR and AF. Death censored any re-intervention free survival over 25 years in TAR and AF. Any re-interventions on the ipsilateral ankle or hindfoot were counted. Re-interventions included any procedure to the ipsilateral ankle and/or hindfoot following the index procedure of interest. Examples of procedures are presented in Figure 5 and Figure 6.

**Figure 5.**
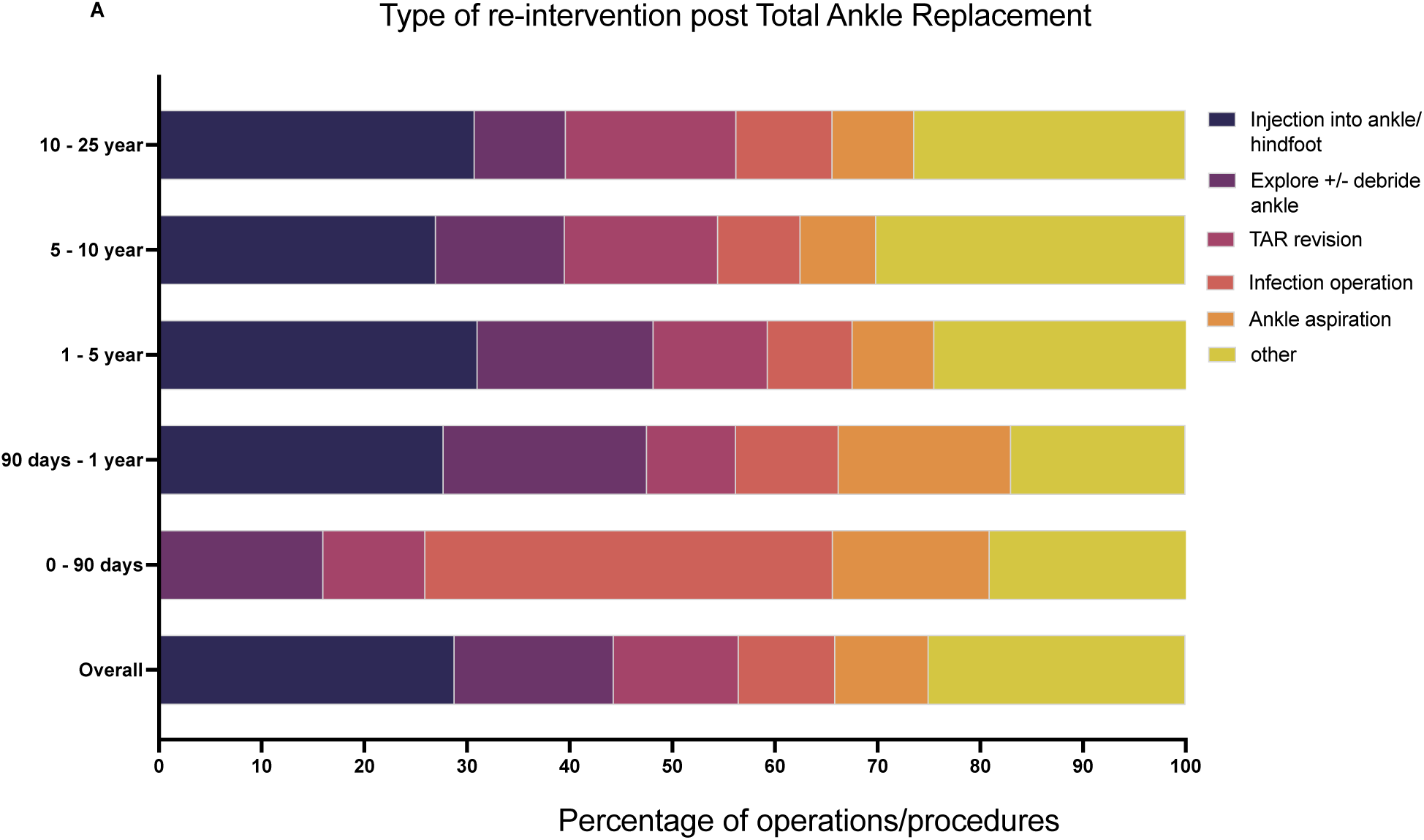
Type and frequency of same ankle/hindfoot re-interventions following index TAR. All operations to the same ankle and/or hindfoot as the original Total Ankle Replacement were analysed and classified based on the nature of the operation. Data shown is the operation as a ***proportion*** of all re-interventions at a particular time point. 0 – 90 days show all operations that occurred between day 0 and day 90. 90 days to 1 year is operations between day 90 and day 365. 1 – 5 years is data between 1 year and up to 5 years post index procedure. 5 – 10 years is operations over 5 years and up to 10 years post index procedure. 10 – 25 years is operations over ten years and up to 25 years post index procedure.

**Figure 6.**
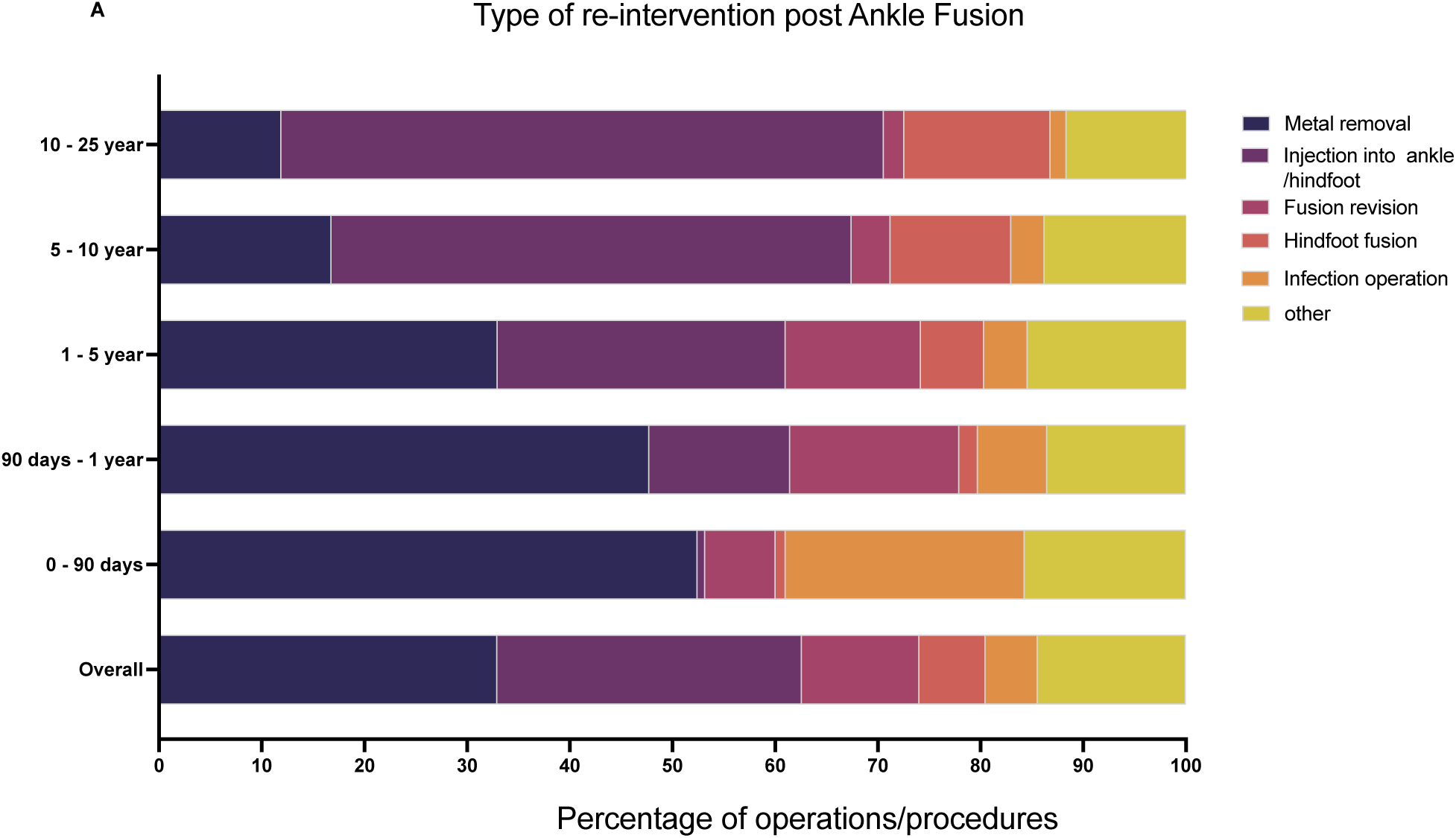
Type and frequency of same ankle/hindfoot re-interventions following index Ankle Fusion. All operations to the same ankle and/or hindfoot as the original Ankle Fusion were analysed and classified based on the nature of the operation. Data shown is the operation as a ***proportion*** of all re-interventions at a particular time point. 0 – 90 days show all operations that occurred between day 0 and day 90. 90 days to 1 year is operations between day 90 and day 365. 1 – 5 years is data between 1 year and up to 5 years post index procedure. 5 – 10 years is operations over 5 years and up to 10 years post index procedure. 10 – 25 years is operations over ten years and up to 25 years post index procedure.

**Table 3.** Re-interventions following Ankle Fusion.

| *Operation* | *Overall* |  | *0 – 90 days* |  | *90 days - 1 year* |  | *1 - 5 years* |  | *5 - 10 years* |  |
| --- | --- | --- | --- | --- | --- | --- | --- | --- | --- | --- |
|  | **N** | **%** | **N** | **%** | **N** | **%** | **N** | **%** | **N** | **%** |
| *Metal removal* | 4147 | 32.96 | 214 | 52.45 | 1368 | 47.75 | 2121 | 33.00 | 322 | 16.80 |
| *Injection into hindfoot* | 3731 | 29.65 | 3 | 0.74 | 393 | 13.72 | 1802 | 28.03 | 971 | 50.65 |
| *Revision fusion* | 1439 | 11.44 | 28 | 6.86 | 472 | 16.47 | 847 | 13.18 | 73 | 3.81 |
| *Hindfoot fusion* | 812 | 6.45 | 4 | 0.98 | 52 | 1.82 | 395 | 6.14 | 225 | 11.74 |
| *Infection* | 639 | 5.08 | 95 | 23.28 | 193 | 6.74 | 273 | 4.25 | 62 | 3.23 |
| *Amputation* | 608 | 4.83 | 19 | 4.66 | 89 | 3.11 | 335 | 5.21 | 123 | 6.42 |
| *Explore +/-debride ankle* | 593 | 4.71 | 25 | 6.13 | 141 | 4.92 | 315 | 4.90 | 77 | 4.02 |
| *Bone cyst graft* | 432 | 3.43 | 6 | 1.47 | 116 | 4.05 | 259 | 4.03 | 39 | 2.03 |
| *Fracture/Fixation* | 15 | 0.12 | 0 | 0.00 | 2 | 0.07 | 8 | 0.12 | 4 | 0.21 |
| *Ankle ligament recon* | 11 | 0.09 | 0 | 0.00 | 0 | 0.00 | 7 | 0.11 | 3 | 0.16 |
| *Tendon operation* | 6 | 0.05 | 0 | 0.00 | 2 | 0.07 | 3 | 0.05 | 0 | 0.00 |
| *Other/misc. codes* | 150 | 1.19 | 14 | 3.34 | 37 | 1.29 | 63 | 0.98 | 18 | 0.94 |
| *Total* | 12583 | 100 | 408 | 100 | 2865 | 100 | 6428 | 100 | 1917 | 100 |

## Discussion

The study is the first national population study of its kind to inform patients and clinicians on the long-term implications of Total Ankle Replacement versus Ankle Fusion for end stage arthritis. Both have been shown to be effective treatments in terms of pain relief, improvements in activities of daily living and patient reported outcome measures^5,8,17,26^. The choice between TAR and AF may have many influences, including the availability of expertise, the age of the patient, cost of treatment, and individual preference (both patient and/or surgeon). Our data demonstrates that both operations are safe, but in the long-term AF is associated with lower revision rates, lower re-operation rates and no increase in the risk of subsequent hindfoot fusion compared with TAR.

### Revision rates of TAR versus AF

The 10 year TAR revision rate observed in this national cohort of 10,335 patients was 10.9% and the 20 year revision rate was 13.5%. This is lower than previously reported in multiple national joint registries. The Norwegian and Swedish registries report TAR revision rates of 32% and 26% respectively at 10 years ^17^. Pooled data of 4,123 patients from three Registries with longer follow-up data (New Zealand, Sweden, and Norway) demonstrated mean revision rates of 34% at 15 years and 38% at 19 years^17^; much higher than our observed 20 year revision rate of 13.5%.

Our observations, however, are consistent with a recent study involving a subset of 5,562 TAR with linked England HES-National Joint Registry data that found ten-year revision rates of 13.8%^27^. It is not clear why TAR revision rates are lower in the United Kingdom compared to more established Registries. Single centre studies have shown low early revision rates for more modern prostheses^26^ and these are increasingly used in England^21^. The NJR-HES linked study also found a survival advantage for the more modern Infinity TAR compared to other implants, but interestingly not when compared to the STAR implant that has been used for decades^18,19^. A major limitation of the HES data is that it does not contain information on the specific implants used. Historical changes in global healthcare provision may also explain some of the observed improvement in TAR survival in more recent Registries, for example the development of orthopaedic sub-specialisation and/or advances in rheumatoid arthritis treatments.

Alternatively, the lower TAR revision rates in England may reflect surgical caution in attempting revision surgery. Revision arthroplasty is technically limited at a global level by the paltry availability of ‘Revision TAR’ implants, and often at a patient level by technical and anatomical challenges (e.g. residual bone stock of the talus). Revision of a TAR to a fusion is therefore a commonly used alternative. An England NJR data linkage study followed 131 patients undergoing revision fusion for failed TAR^28^. Successful fusion was inferred by a ‘single stage’ procedure in 79% of patients, while 21% required a second stage. The 5-year ‘survival’ of revision ankle fusion was 72%. The mean follow-up period was just under 4 years and the national population database was unable to differentiate between fixation methods. There is considerable technical variability of a ‘TAR Revision to fusion’ procedure ranging from an ankle fusion to a highly complex ankle/hindfoot reconstruction involving allografting of bone defects and fusion of the tibio-talo-calcaneal (TTC) complex. This range of operative complexity, the associated morbidity and requisite rehabilitation are not at all represented in our study. Pfahl et al. were able to compare the outcomes of revision ankle arthroplasty, revision to ankle fusion and tibio-talo-calcaneal fusion of 111 patients with failed TAR^29^. In their cohort, only 4 of 46 revision to ankle fusions went on to a non-union requiring further surgery. None of the patients that underwent tibio-talo-calcaneal fusion failed (defined as requiring further surgery) and there was a trend for improved postoperative patient reported outcome measures (PROMS) compared with revision to ankle fusion and revision ankle arthroplasty. Other single centre studies of TAR revision to fusion report similar non-union rates of 20%.

TAR revision procedures are complex, time consuming, require a high level of experience (that is difficult to achieve given the paucity of procedures), and have poor outcomes compared to primary TAR ^24,29^. Collectively, the above are all reasons why HES revision rates of TAR are not necessarily the same as failure rates of TAR. It is possible there is a large cohort of patients currently living with symptomatic and failing TAR that are being dissuaded from surgery. The large number of observed minor operations performed enroute to a far more complex TAR revision operation supports this understandable procrastination (Figure 5 and Table 3). 52% of all re-interventions post TAR in England involve ankle exploration/debridement, aspiration, and/or injection.

The Swedish Arthroplasty Register report of 118 fusions for failed TAR found that post revision functional scores were poor and less than half of patients were satisfied despite successfully achieving fusion in 90% of patients^23^. Our own single centre study used the MOxFQ^30^ to assess postoperative patient reported outcomes and the scores were similarly poor^31^. In particular, patients that received either a TTC fusion or revision to ankle fusion using an intra-medullary nail had mean index scores of 40.7 and 35.2 out of 64 respectively (where a lower score indicates better outcomes). To put these results in context, primary ankle fusion was previously shown to reduce the mean MOxFQ score from 53.8 to 22.9 and, in the case of big toe fusion, from 43.0 (preoperatively) to 12.1^32^. Yet despite relatively poor postoperative MOxFQ scores, 73% of patients would recommend the revision surgery to their friend/family member and 73% reported an overall improvement, suggesting preoperative symptoms were even worse.

Without pre- and post-operative PROMS, we can only hypothesise that these collective observations are indicative of a high operative ‘threshold’ preventing a larger number of these technically challenging cases being performed annually. A similar phenomenon has previously been described in other areas of arthroplasty. Most notably, the preoperative Oxford Knee Score threshold is higher in patients undergoing revision of a total knee replacement versus uni-compartmental knee replacement; a failing total knee replacement needs to be that much more debilitating before the (more complex) revision is offered^33^.

### Low risk of adjacent joint fusion post AF and TAR

Our study does not support the hypothesis that TAR is better than AF at reducing the requirement for adjacent joint fusion. A major concern of AF has been that sacrificing ankle range of motion for pain resolution might come with the long-term ‘cost’ of adjacent joint fusion and an eventual rigid ankle hindfoot fusion ^13,14^. SooHoo et al. found TAR was associated with a lower rate of subtalar joint (STJ) fusion at five years versus AF; 0.7% versus 2.8% respectively (hazard ratio = 0.28, 95% confidence interval; 0.09 to 0.87, p = 0.03)^14^. In a recent single centre study of over 250 patients undergoing arthroscopic AF, only 4% (11 patients in total) required a STJ fusion^13^. Nonetheless Kaplan-Meier analysis estimated a STJ ‘survivorship’ of only 74% at 9 years post AF, highlighting the potential limitations of survival analysis when applied to smaller cohorts. In this 25-year study of over 10,335 TAR and 30,704 AF, we defined hindfoot fusion very broadly and included any codes for subtalar joint fusion, triple fusion, hindfoot fusion and talo-navicular joint fusion. Based on this national population data, the risk of subsequent hindfoot fusion is very low and not significantly different to the hindfoot fusion rate post TAR. The latter also excludes patients that received a revision TAR, many of whom underwent a TTC fusion as their ‘revision TAR’ procedure and ended up with a rigid ankle/hindfoot complex. It also excludes any patients who had a primary TAR following a previous hindfoot fusion (Supplementary Figure 2).

Just as there may be an unwillingness to revise a TAR, there may also be resistance to fusing the hindfoot in someone with an AF. A limitation of this study is that it does not investigate the presence of symptomatic hindfoot arthritis, only the rates of hindfoot fusion. A further limitation of the HES database is that it does not include functional data or patient reported outcome measures (PROMS). The TARVA prospective RCT found significant, and similar, improvements in Manchester Oxford Foot Ankle Questionnaire (MOXFQ) at 1 year following both TAR and AF^5^.

In the long-term our observations do not support the hypothesis that TAR is protective of ankle-hindfoot function or survivorship compared to AF.

### Mortality and morbidity

Both TAR and AF are safe operations and the associated 90-day mortality and morbidity rates are low. The 90-day mortality rate post AF was 0.41 versus 0.23 post TAR (OR 0.57, Figure 1). Beyond 7 years after TAR, the observed mortality rate is greater than in patients post AF (Supplementary Figure 3). The survival curves continue to diverge such that by 15-years, 59% (95% CI 58%-61%) of TAR patients have died compared to 67% (95% CI 66% - 68%) of AF patients. TAR were more commonly performed on patients over the age of 60 years old (78% versus 54% of AF, Table 1), although this alone may not fully explain the observed differences in patient survival. This is not a matched study, however the two groups were similar in terms of other demographics, index of social deprivation and Charlson co-morbidity indices (Table 1). Future work involving well matched cohorts is required to investigate if there is a genuine associated long-term reduction in survival associated with TAR.

The risks of serious adverse effects post TAR and AF are low. The observed pulmonary embolus rate post AF of 0.58% is higher than TAR (0.23%), but both are low overall in comparison to recently published prospective UK Foot Ankle Thrombo-Embolism Audit (UK-FATE) data showing an overall Foot Ankle peri-operative VTE rate of 1.1% ^34^. A meta-analysis in 2017 of 6 studies found an increased rate of complications post AF compared to TAR but a lower re-intervention and revision rate ^35^. The Canadian Orthopaedic Foot and Ankle Society (COFAS) Prospective Ankle Reconstruction Database of 388 ankles (281 TAR and 107 AF) found higher peri-operative complications following TAR, and similar improvements in outcome scores^36^. A more recent meta-analysis involving 13 studies found no difference in post operative complication rates between TAR and AF^37^. Interestingly, it also found no difference in the Short Form-36 scores between the TAR and AF groups but improved Foot and Ankle Ability Measure scores in the TAR group.

### Limitations

All HES database studies are limited by the accuracy of the recorded OPCS-4 and ICD-10 codes. The results and conclusions drawn are susceptible to misclassification or omission of codes and also do not include patients that had incomplete data (Supplementary Figures 1 and 2).

A major limitation of this study is the confounding effect of the inherent selection bias of patients receiving either AF or TAR. We chose to analyse un-matched population data rather than apply adjustment models, for example propensity score matching, that greatly diminishes the numbers in each group^38^. We began with the assumption that for the last 25 years, healthcare professionals and patients have used the information available to them to make the most appropriate individual choice of TAR or AF. We were interested in the real world long consequences of these choices. The patient demographics are not significantly different with the notable exception of age, although estimate of effect testing reveals this difference to be of minimal practical significance (Table 1). This fits with common clinical practice whereby younger, fitter, more active patients are thought to have a greater risk of TAR revision and so are more likely to undergo AF. The concern has been that they are then more likely so require further hindfoot fusion surgery, but our results do not support this.

Further work will focus on subgroup analysis of open versus arthroscopic ankle fusion as well as an economic analysis of TAR versus AF. Can we justify the additional implant cost of TAR over AF based on this long-term data?

## Conclusions

Both Total Ankle Replacement and Ankle Fusion for end-stage arthritis are safe operations. The observed revision rate of TAR in this national population study is 13.5% over 20 years and lower than reported previously. There was no long-term difference in survivorship of adjacent joints in AF versus TAR. Overall, these observations alone do not support the use of TAR over AF in treating end stage isolated ankle arthritis.

## Data Availability

All data produced in the present work are contained in the manuscrip

---

**Supplementary Figure 1.**
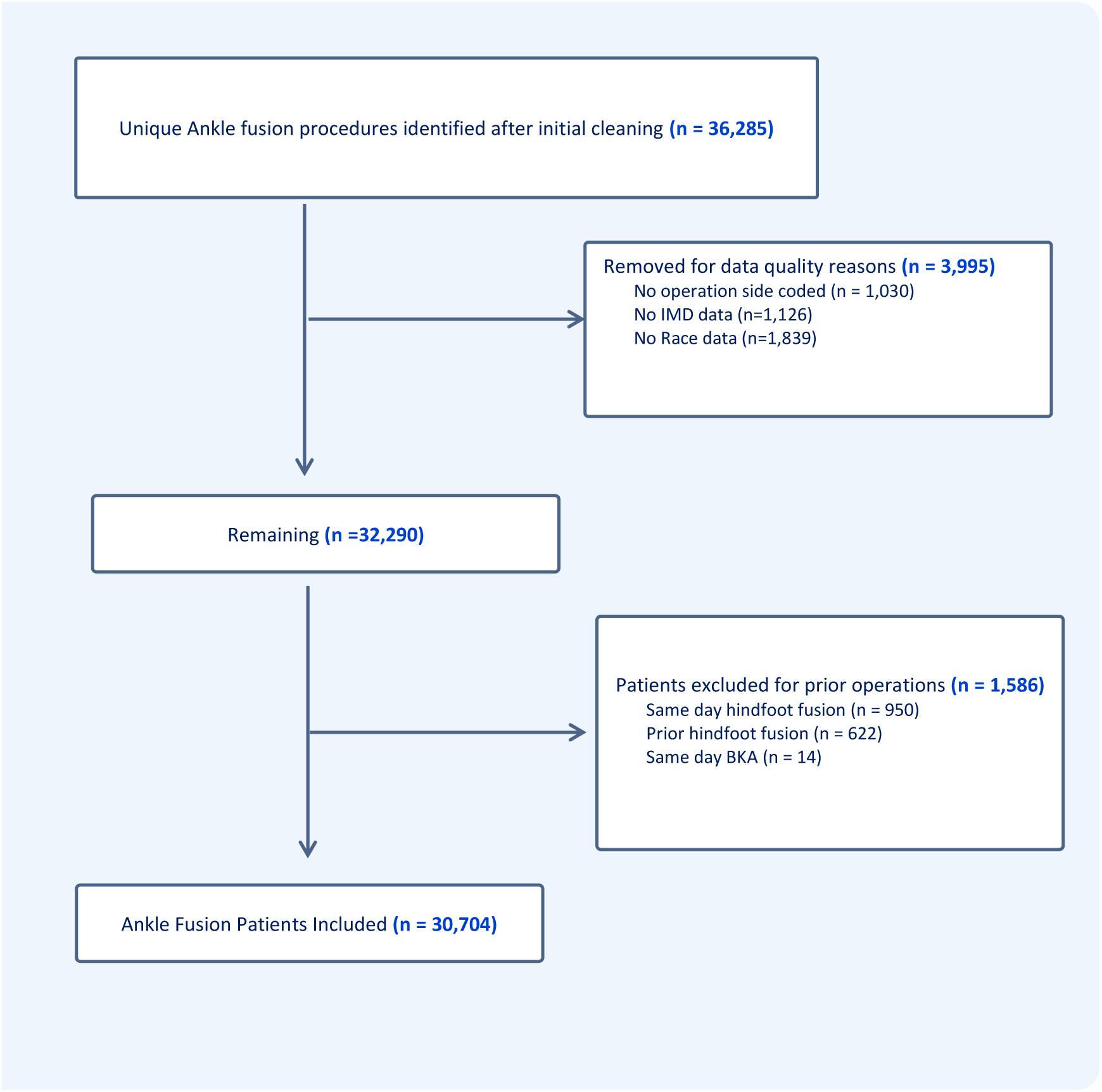
Ankle fusion procedure selection diagram. The above figure outlines additional steps beyond the primary data cleaning process to ensure data quality in the cases used for analysis. Data with no operation site coding was excluded. The absence of race information or index of multiple deprivation data (IMD) also resulted in exclusion of cases. Finally, patients who had prior or same day hindfoot operations were excluded from analysis as they would act as a confounder in the analysis of hindfoot disease progression.

**Supplementary Figure 2.**
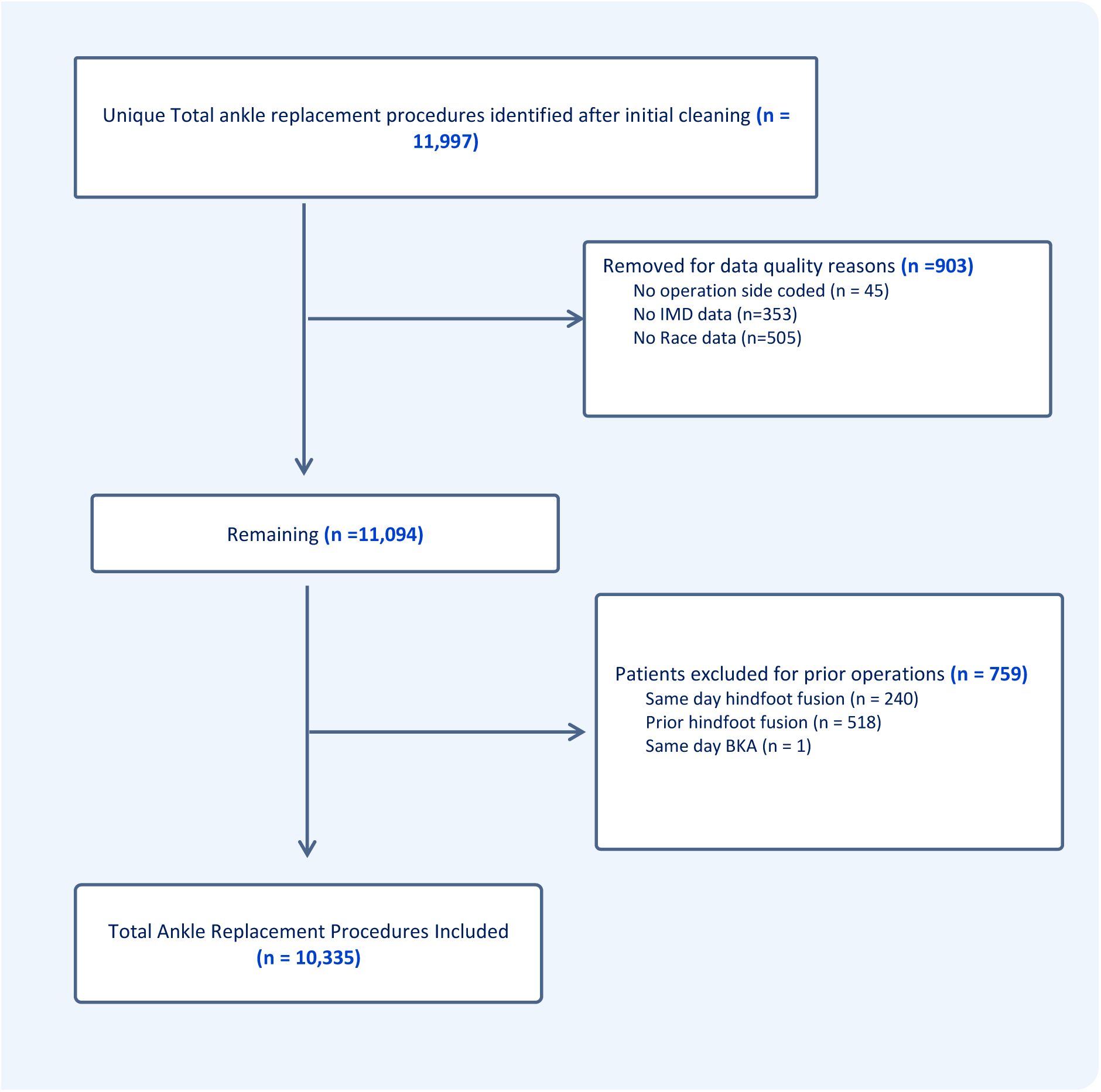
Total ankle replacement procedure selection diagram. The above figure outlines additional steps beyond the primary data cleaning process to ensure data quality in the cases used for analysis. Data with no operation site coding was excluded. The absence of race information or index of multiple deprivation data (IMD) also resulted in exclusion of cases. Finally, patients who had prior or same day hindfoot operations were excluded from analysis as they would act as a confounder in the analysis of hindfoot disease progression.

**Supplementary Figure 3.**
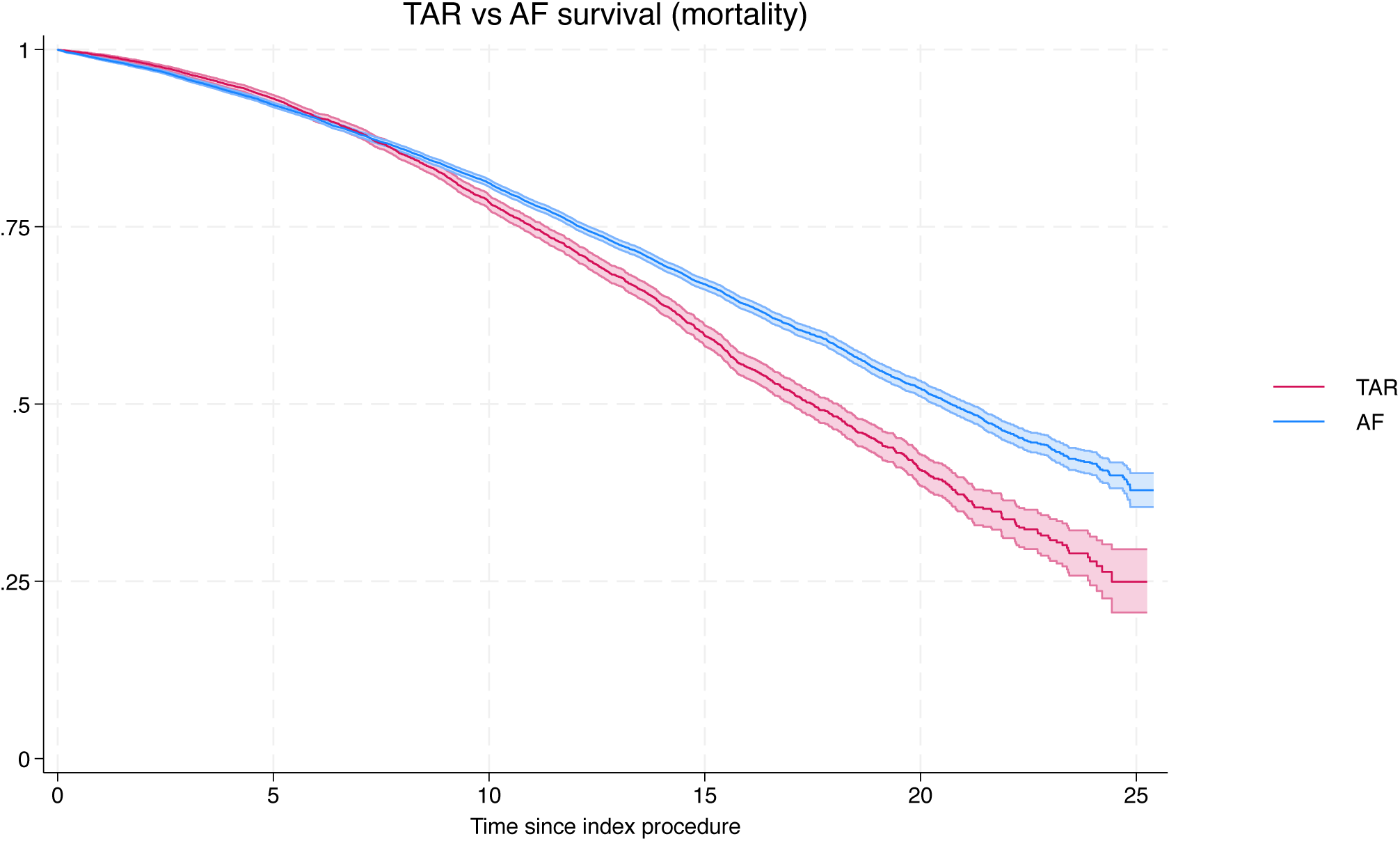
25-year mortality in TAR vs AF patients.

**Supplementary Table 2.** Unadjusted and adjusted hazard ratios of revision following TAR or AF. Data shown is hazard ratio +/- 95% CI. Adjusted HR are adjusted for index procedure, sex, age, race, IMD quintile and Charlson score.

|  | **Unadjusted HR** |  | **Adjusted HR** |  |
| --- | --- | --- | --- | --- |
|  | HR | 95% CI | HR | 95% CI |
| **Index Procedure** |  |  |  |  |
| AF | 1 |  | 1 |  |
| TAR | 2.93 | (2.63-3.27) | 3.28 | (2.92-3.67) |
| **Sex** |  |  |  |  |
| Male | 1 |  | 1 |  |
| Female | 0.91 | (0.81-1.02) | 0.84 | (0.75-0.94) |
| **Age group** |  |  |  |  |
| 0 - 19 | 0.55 | (0.25-1.19) | 0.57 | (0.26-1.22) |
| 20 - 39 | 1 |  | 1 |  |
| 40 - 59 | 1.20 | (0.98-1.48) | 0.93 | (0.75-1.15) |
| 60 - 79 | 1.01 | (0.82-1.24) | 0.58 | (0.46-0.72) |
| 80+ | 0.44 | (0.29-0.67) | 0.23 | (0.15-0.35) |
| **Race** |  |  |  |  |
| White | 1 |  | 1 |  |
| Black | 0.09 | (0.013-0.66) | 0.09 | (0.01-0.7) |
| Asian | 0.84 | (0.48-1.45) | 0.85 | (0.49-1.48) |
| Mixed | 0.77 | (0.19-3.10) | 0.84 | (0.21-3.38) |
| Other | 0.66 | (0.21-2.05) | 0.78 | (0.25-2.43) |
| **IMD** |  |  |  |  |
| Most deprived 0-20% | 1.22 | (1.03-1.43) | 1.20 | (1.02-1.41) |
| More deprived 20-40% | 1.20 | (0.87-1.21) | 1.02 | (0.86-1.21) |
| Middle 40-60% | 1 |  | 1 |  |
| less deprived 60-80% | 0.97 | (0.82-1.15) | 0.98 | (0.81-1.16) |
| least deprived 80-100% | 0.94 | (0.78-1.13) | 0.97 | (0.81-1.16) |
| **Charlson score** |  |  |  |  |
| Mild (1-15) | 1 |  | 1 |  |
| Moderate (16-30) | 1.34 | (1.16-1.55) | 1.25 | (1.09-1.45) |
| Severe (31-50) | 1.72 | (1.52-1.95) | 1.74 | (1.54-1.98) |

**Supplementary Figure 4.**
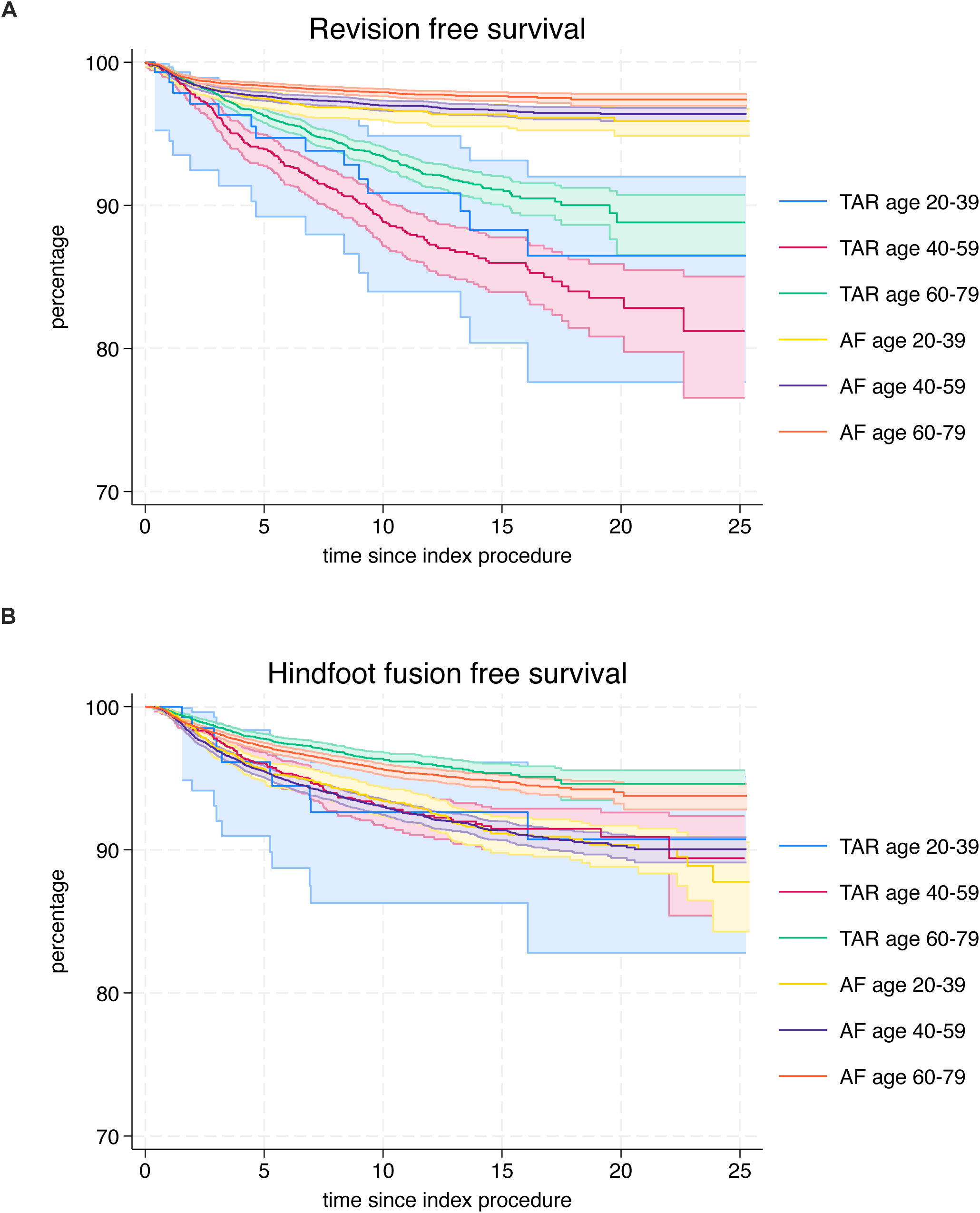
KM plots of revision free survival (A) and hindfoot fusion free survival (B) in both TAR and AF patients. The 0-19 and 80+ age groups were supressed due to small numbers.

## Notes

Conflicts of Interest: There are no conflicts of interest for the authors.

### Competing Interest Statement

The authors have declared no competing interest.

### Funding Statement

This study was funded by RCS

